# The voices of patients and caregivers – a qualitative interview study on what influences levels of mobility, among patients hospitalized following hip fracture surgery

**DOI:** 10.64898/2026.07.03.26357215

**Authors:** Sofie Tscherning Lindholm, Kira Marie Skibdal, Thomas Bandholm, Mette Merete Pedersen, Jeanette Wassar Kirk, Maria Swennergren Hansen

## Abstract

**Purpose:** To explore patient and caregiver perspectives on factors influencing mobility during hospitalization after hip fracture surgery, and how these are experienced and negotiated in everyday hospital practice.

**Materials and methods:** A qualitative interview study informed by a hermeneutic-phenomenological perspective was conducted in a hospital setting in Denmark. Using purposive sampling with maximum variation, ten patients and nine caregivers were interviewed during hospitalization. Data were analyzed using reflexive thematic analysis following Braun and Clarke.

**Results:** Five interrelated themes were identified; (1) Body and mind in transition; (2) Communication as a prerequisite for safety and mobility; (3) Structural barriers and ambiguities in responsibility; (4) The physical environment and ward culture; and (5) Mobility as preparation for life after discharge. Across themes, mobility emerged as a socially shaped and negotiated practice through everyday interactions, communication, organizational routines, and situational support during hospitalization.

**Conclusions:** Mobility during hospitalization after hip fracture surgery emerged as a context-dependent and socially shaped practice rather than a purely physical task. These findings suggest that rehabilitation during hospitalization may need to attend not only to mobility prescription, but also to relational, communicative, and contextual aspects of everyday ward routines that shape patients’ confidence and participation.

**Trial registration:** OSF Registration DOI: 10.17605/OSF.IO/XT76H

## INTRODUCTION

Hip fracture is one of the most common and severe injuries among older adults and is associated with high morbidity (1), mortality (2), and loss of independence (3). Beyond these clinical and epidemiological consequences, hip fracture often represents a profound disruption in older people’s everyday lives, identities, and relationships, where mobility becomes closely intertwined with vulnerability, fear of dependency, and hope for recovery (4–9). Patients describe recovery as an existential and relational journey, in which mobility is experienced as both emotionally demanding and a way of restoring autonomy and control (4–6,9,10). From a hermeneutic-phenomenological perspective, mobility after hip fracture can therefore be understood not only as a measurable physical outcome, but as a lived and socially negotiated practice through which patients and caregivers make sense of the bodily vulnerability, safety, and possibilities for the future (5,10–13). In Denmark, approximately 7,000 individuals sustain a hip fracture annually (14), and the incidence is expected to increase in line with population ageing (15,16). Despite advances in surgical and perioperative care, recovery trajectories remain poor, with only about half of patients regaining their pre-fracture level of mobility (17–19). Hip fracture is thus considered both a major personal event with significant impact on the patients’ health-related quality of life (4,6,7) and a significant societal challenge due to prolonged rehabilitation (20), increased healthcare costs (21), and a high risk of institutionalization (18).

Mobility is a cornerstone of post-operative care following hip fracture surgery, where movement in all forms, assisted or independent, plays a central role in recovery (18,22). Clinical guidelines and enhanced recovery programs consistently recommend early mobility (23,24), and getting patients out of bed and move on the day of or the day after surgery has been shown to reduce complications, mortality, and length of hospital stay (21,25). Despite clear recommendations, observational studies show that many patients remain physically inactive during hospitalization, spending most of their time in bed or seated (26,27). Barriers to mobility include pain (4,10,28), fatigue (4,10), fear of falling (4,29), comorbidities (18,30), and restricted weight-bearing instructions (25,31), as well as structural and organizational challenges such as staff shortages (32) and unclear allocation of responsibilities (32,33). Although clinical guidelines for mobility and rehabilitation after hip fracture are well documented (18,23), their implementation in everyday hospital practice remains inconsistent (28,32,34). This suggests that factors beyond clinical evidence influence how mobility-related activities are initiated, supported, and sustained during hospitalization.

Qualitative research highlights that mobility after hip fracture is not only a physical task but also a deeply personal and relational process (5,10). Patients describe the recovery journey as shaped by both vulnerability and hope, where communication, support, and meaningful engagement with healthcare professionals play crucial roles in regaining confidence and autonomy (4,5,10). Caregivers often become important facilitators of mobility and recovery and providers of emotional support, yet report feeling unprepared, under-informed, and burdened by responsibility (35,36). These perspectives suggest that successful rehabilitation requires not only clinical expertise but also a patient- and caregiver-centered approach that addresses both physical and psychosocial needs.

There is a growing call for interventions that integrate patient, caregiver, and staff perspectives to overcome barriers and promote sustainable mobility practices across hospital and rehabilitation settings (32,37). However, existing qualitative studies have predominantly explored patient and caregiver experiences after discharge or during later phases (36,38), with relatively limited attention to how mobility is experienced and negotiated during the acute hospital stay (35). While mobility is often addressed as a physical and clinical outcome (e.g., walking ability, or balance) (25,39), this perspective may overlook how mobility is embedded in everyday social practices shaped by interactions, communication, and organizational routines. Understanding how mobility is experienced and enacted in context is therefore essential to inform rehabilitation practices. To complement existing staff-centered research on mobility practices after hip fracture (32–34) and inform more person-centered mobility practices, this study aimed to explore patient and caregiver perspectives on factors influencing mobility during acute hospitalization after hip fracture surgery, and how mobility is experienced and negotiated in everyday hospital practice.

## MATERIALS AND METHODS

### Study design

A qualitative design was chosen to gain in-depth understanding of patients’ and caregivers’ lived experiences of mobility during hospitalization (40). The study was informed by a hermeneutic-phenomenological perspective inspired by Gadamer (11,12), emphasizing that understanding emerges through interpretation and dialogue between participants’ narratives and researchers’ pre-understandings (13). This perspective guided the study by acknowledging that meanings are not simply discovered in the data but co-constructed through an iterative interpretive process.

Data were analyzed using reflexive thematic analysis as described by Braun and Clarke (41). In this approach, themes are generated through active and reflective engagement with the data, and researcher reflexivity is considered central to knowledge production. Rather than aiming for coding consensus or objectivity, reflexive thematic analysis recognizes the researcher’s central interpretive role and understands themes as patterns of shared meaning that are actively constructed through reflexive engagement with the data (42,43).

Within this approach, hermeneutic phenomenology informed the interpretive engagement with participants’ lived experiences, while reflexive thematic analysis provided a flexible analytic framework for identifying patterns of shared meaning across participants. Reflexive thematic analysis was chosen over interpretative phenomenological analysis (44) and other phenomenological approaches (45,46), as the aim was not to prioritize in-depth analysis of individual cases, but to develop an interpretive understanding of patterns of shared meaning across participants.

Combining hermeneutic phenomenology with reflexive thematic analysis enabled an exploration of participants’ lived experiences while situating them within broader patterns of meaning across the dataset.

### Study setting

This study was conducted at a university hospital located in the Capital Region of Denmark. Data were collected from the hospital’s orthopedic department, which encompassed two units accommodating up to 32 patients in one unit and 26 in the other. The healthcare professionals included nurses, nurse assistants, orthopedic surgeons, and pharmacists. Rehabilitation services, physiotherapy and occupational therapy, were centrally organized. Physiotherapy services were available daily, including weekends, although the frequency and duration of sessions varied across weekdays and weekends. Physiotherapists were generally the same individuals each day, ensuring continuity of care. In contrast, occupational therapy services varied more. The service was delivered by a team of six therapists covering the entire hospital and engaging in teaching and development tasks, which resulted in less consistency in staff and treatment at the orthopedic department.

### Researcher characteristics and reflexivity

The research group consisted of six members (STL, KS, TB, MMP, JWK, and MSH) with diverse academic achievements (e.g., master’s degree or PhD degree), and clinical backgrounds (e.g., physiotherapists and nurses). All researchers were part of the HIP-ME-UP research program and had prior experience in conducting qualitative interview studies. The group shared the preconception that implementing evidence-based knowledge in clinical practice is inherently challenging. These preconceptions were actively discussed throughout data collection and analysis to remain attentive to participants’ own priorities and meanings. STL, who conducted all interviews, had a clinical background in rehabilitation but no therapeutic relationship with participants, positioning her as both an insider to the clinical context and an outsider to participants’ individual care trajectories.

### Trustworthiness

Trustworthiness was addressed through strategies enhancing credibility, dependability, transferability, and confirmability (47,48). Credibility was supported by researchers reflecting on their preconceptions based on their clinical backgrounds as healthcare professionals, maintaining openness to participants’ narratives and avoiding leading questions during interviews. Follow-up and iterative questions, along with reflective notes written during or immediately after the interviews, supported accurate interpretation. Credibility was further strengthened by presenting both consistent patterns and divergent views across interviews, and by relating findings to existing literature and theoretical frameworks. Patients contributed to the development of the interview guide (n=3) and participated in test interviews (n=1), leading to minor revisions in wording and sequencing of questions to improve clarity, relevance, and comprehensibility from a patient perspective. Dependability was addressed by having the same researcher (STL) conduct all interviews using a semi-structured interview guide, with three interviews observed by MSH to provide feedback on the interview technique. All six authors were involved in the analysis and interpretation including a thorough discussion of categories. Transferability was supported by providing rich descriptions of context, participants, and findings. Confirmability was pursued through the involvement of six researchers in the data analysis. Researcher characteristics and potential biases were made explicit and actively considered to support reflexivity. The findings were linked to empirical data and illustrated through participant quotations labelled with unique codes. Also, the rationale for the methods and procedures was clearly outlined.

### Participants

The study included patients who had undergone hip fracture surgery and caregivers (informal caregivers, e.g., relatives) of patients hospitalized in the department. Eligible patients were aged 60 years or older, able to provide written informed consent and had a pre-fracture Cumulated Ambulation Score (CAS) (49) of ≥ 3 points (as recalled), reflecting their degree of independence in basic mobility (e.g., getting in and out of bed) (score range 0–6, higher scores indicate greater independence). Patients were excluded if they had a terminal illness, delirium, or cognitive impairment. Caregivers of eligible patients were included regardless of the patient’s cognitive status. The caregivers were not necessarily related to the interviewed patients but were relatives of patients hospitalized in the department.

Eligible participants were identified through a maximum variation purposive sampling, a prevalent technique in qualitative research (50). This approach involves selecting individuals with specialized knowledge or direct experience related to the phenomenon under investigation. Emphasizing maximum sampling variation, the strategy seeks to capture diverse perspectives, thereby elucidating common patterns inherent in the variations (40). Physiotherapists assisted in identifying eligible patients based on the inclusion criteria and STL approached them during their postoperative hospitalization, when deemed clinically stable by ward staff. The target sample size was assessed based on the concept of information power, which was influenced by the study aim, sample specificity, theoretical framework, anticipated quality of dialogue, and planned type of analysis (51). We considered our study aim to be focused yet sufficiently broad, the sample specificity to be dense, no predefined theoretical framework informing sampling, the quality of dialogue to be strong, and the analysis strategy to be cross-case. Therefore, we pre-defined the number of participants as ten patients and ten caregivers in total in the department.

### Data collection

Semi-structured individual interviews were conducted consecutively by the first author prior to patient discharge. Most interviews took place in-person in private rooms at the hospital ward during patient hospitalization, while three interviews with caregivers were conducted by telephone due to logistical constraints. The telephone format was chosen in accordance with participants’ preferences and did not appear to compromise the depth or richness of the data, as reflected in the length and content of the interviews. Prior to the interviews, mobility (52) was introduced as an umbrella concept encompassing mobilization (53,54), physical activity (55), and exercise therapy (56). However, during the interviews, participants primarily referred to “getting out of bed and moving,” reflecting a more integrated and experience-based understanding of mobility. Accordingly, mobility was retained as the central analytic concept. Reflexivity was supported through memo writing after each interview and regular discussions among authors to reflect on preconceptions and maintain transparency during analysis.

An interview guide was employed during the interviews, developed based on previous studies from HIP-ME-UP research program (26,57,58) and existing evidence (33,59–61). It was also adapted to the specific characteristics of each participant group - patients and caregivers. The interview guide covered themes such as experiences with mobility during hospitalization, communication regarding mobility with healthcare professionals, perceived barriers and facilitators to mobility, and expectations for discharge and recovery. The interview guide included a total of 17 questions, e.g., “What do you consider to be important reasons for getting out of bed?”, “What kind of movement do you think is best for you while you are in the hospital – for example getting out of bed, being physically active, or doing exercises?”, and “Who do you feel helps ensure that you get out of bed and move around during your hospital stay?”. All interviews were audio-recorded with an Olympus WS-853 digital voice recorder and lasted between 20 and 50 minutes. Field notes were made immediately after each interview to capture contextual details and reflections relevant to interpretation. Transcription of the interviews was guided by a standardized transcription protocol, transcribed using the Viceron application (62), and the software program NVivo (Version 14) was employed to organize and analyze the qualitative data (63).

In cases of potential language barriers, where participants spoke a language other than Danish or English, a professional interpreter was prepared to assist during the interview; however, all participants in this study spoke Danish, and interpreter services were not required.

Transcripts were not returned to participants for comment due to the acute hospital setting and practical reasons. However, interpretations were discussed iteratively within the multidisciplinary research group to ensure credibility.

### Data analysis

Demographic data were collected and managed using Research Electronic Data Capture (REDCap) (64).

The interview data were analyzed using reflexive thematic analysis as described by Braun and Clarke (41,42), following iterative phases (41). The analysis involved movement between individual accounts and the whole dataset, where interpretations were continuously questioned and refined in light of emerging patterns and researchers’ pre-understandings. Although the analysis was primarily inductive, coding was grounded in participants’ narratives while informed by the researchers’ clinical and academic backgrounds, which were continuously reflected upon.

First, STL and MSH familiarized themselves in the data through repeated reading of all 19 interview transcripts. Subsequently, STL conducted systematic line-by-line coding in NVivo, identifying meaningful segments related to mobility. Coding was approached as an analytical process, attending to both semantic content and underlying meanings. Codes were examined for patterns of shared meaning and clustered into preliminary themes by STL, JWK, and MSH. The preliminary themes were presented to the multidisciplinary research group (STL, KS, TB, MMP, JWK, and MSH), where they were discussed and critically examined in a series of analytic meetings. The broader research group engaged in iterative discussions to refine themes, focusing on analytic depth rather than coding consensus. Through ongoing reflexive dialogue, themes were refined to ensure internal coherence, clear distinction, and grounding in the empirical material. Finally, a coherent analytic narrative was developed, integrating descriptive accounts with interpretive insights and illustrated using participant quotations labeled with unique identifiers.

### Ethics

Before the interviews, participants were given verbal and written information about the study. Written informed consent was obtained for participation, audio-recording, secure storage, analysis, and publication of interview data. Participants also consented to the researchers accessing relevant information from medical records, including operation type, and pre-fracture and current mobility status (Cumulated Ambulation Score, (49)), to contextualize interview data. All data were stored on the hospital’s encrypted research server. Identifying information was removed during transcription, and each participant was assigned a unique study ID for data management. To protect participant anonymity in this manuscript, all names are pseudonyms and potentially identifying information, including exact ages, has been generalized throughout.

To minimize potential power imbalances in the acute hospital setting, recruitment and informed consent were conducted solely by the researcher and not by treating healthcare professionals. Participants were explicitly informed that declining participation would have no consequences for their treatment or care and that they could withdraw at any time without justification.

Patients were approached only after confirmation of medical stability and were given time to consider participation. Because interviews were conducted during acute hospitalization, particular attention was paid to patients’ physical and emotional condition. Interviews were scheduled to avoid interference with treatment or care routines and were adapted to patients’ energy levels and preferences. Interviews took place in private and quiet locations to ensure confidentiality and minimize disturbance. At the beginning of each interview, efforts were made to establish a respectful and supportive atmosphere, and patients were encouraged to speak freely and take breaks if needed.

Caregivers were approached opportunistically when present at the department. In addition, written invitation materials were left at patients’ bedside to allow caregivers to contact the researcher if interested in participation. Interviews with caregivers were arranged flexibly according to their preferences regarding time and location, including telephone interviews when appropriate.

Informed consent materials were available in multiple languages (Arabic, Somali, Turkish, Urdu/Pakistani, and English) in addition to Danish, but were not utilized due to recruitment limitations.

The study received ethical approval from the Scientific Ethical Committee for the Capital Region in Denmark (F-24041115) and was registered in Privacy, the research inventory of the Capital Region of Denmark (p-2024-16885). The study was conducted in accordance with the Declaration of Helsinki (65).

### Reporting standards

The study was reported in adherence with the Consolidated Criteria for Reporting Qualitative Research (COREQ) checklist to ensure comprehensive and transparent reporting of the research process and findings (66). The overarching project was pre-registered at OSF (https://doi.org/10.17605/OSF.IO/XT76H), including the overall research aims, sampling strategy, and an initial analytic approach. As the registration covered multiple related sub-studies, the present analysis represents one component of the broader protocol. Data from healthcare professional interviews have been reported separately (58), whereas the current manuscript reports findings from patient and caregiver interviews.

The initial protocol specified an inductive content analysis; however, during familiarization with the data, the analytic approach was revised to reflexive thematic analysis. This change was made to better align with the study’s hermeneutic-phenomenological orientation and to support an interpretive analysis of patterns of shared meaning across participants. The deviation from the pre-registered analytic plan is reported transparently. The analysis remained consistent with the overall methodological framework described in the registration while allowing flexibility in line with qualitative research principles (42,43).

## RESULTS

We recruited ten patients (table 1) and nine caregivers (table 2). Patients were male (n=3) and female (n=7), aged 69-89 years (mean age=80), with educational backgrounds ranging from less than ten years (n=3), between 10-12 years (n=2), to more than 12 years of education (n=5). Caregivers were male (n=3) and female (n=6), aged 31-82 years (mean=60), with educational background in the range of less than ten years (n=1), between 10-12 years (n=1), to more than 12 years of education (n=7). Caregivers were spouses (n=2), children or in-laws (n=5) or grandchildren (n=2).

**Table 1.**
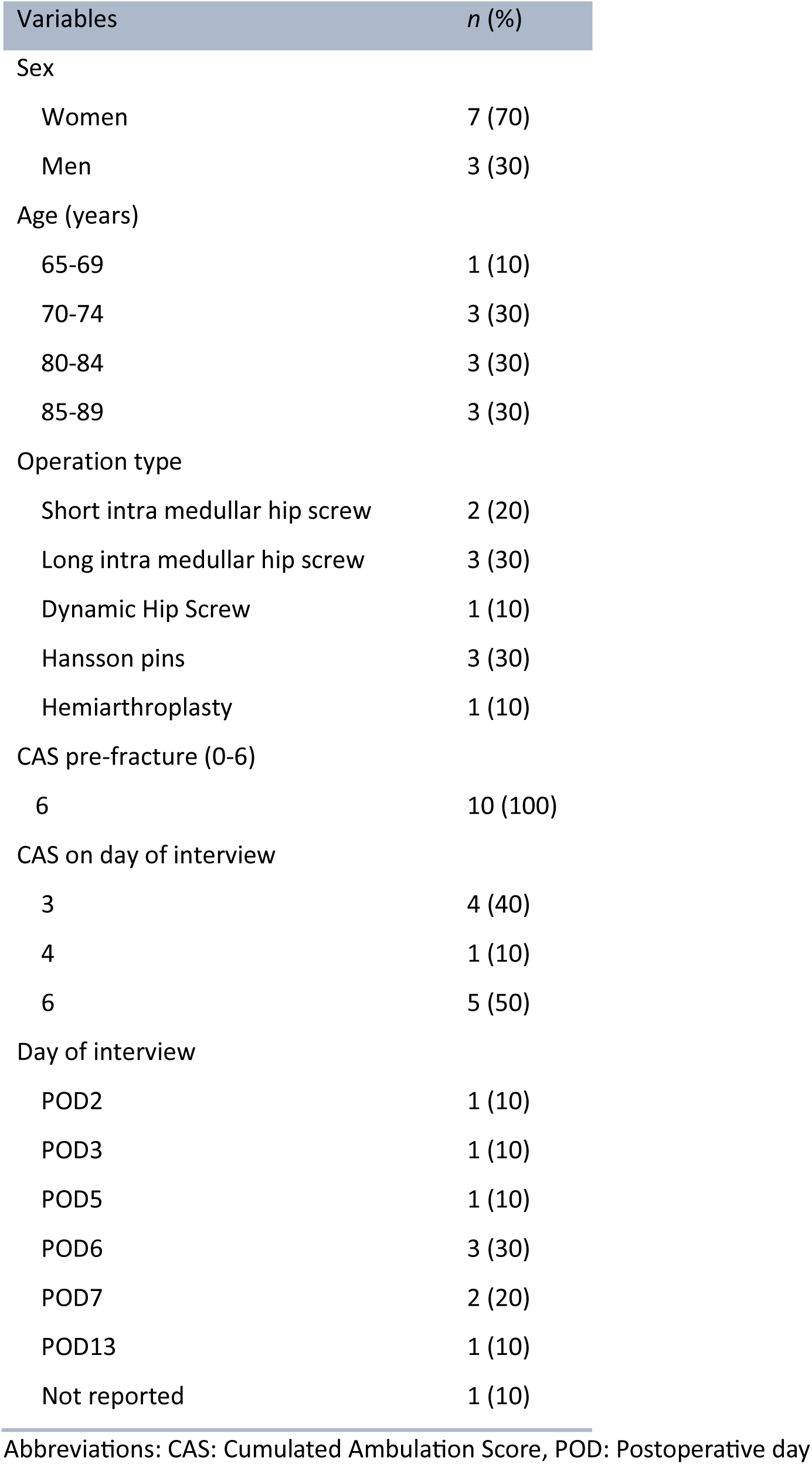
Description of patients (n=10).

| Variables | *n* (%) |
| --- | --- |
| Sex |  |
| Women | 7 (70) |
| Men | 3 (30) |
| Age (years) |  |
| 65-69 | 1 (10) |
| 70-74 | 3 (30) |
| 80-84 | 3 (30) |
| 85-89 | 3 (30) |
| Operation type |  |
| Short intra medullar hip screw | 2 (20) |
| Long intra medullar hip screw | 3 (30) |
| Dynamic Hip Screw | 1 (10) |
| Hansson pins | 3 (30) |
| Hemiarthroplasty | 1 (10) |
| CAS pre-fracture (0-6) |  |
| 6 | 10 (100) |
| CAS on day of interview |  |
| 3 | 4 (40) |
| 4 | 1 (10) |
| 6 | 5 (50) |
| Day of interview |  |
| POD2 | 1 (10) |
| POD3 | 1 (10) |
| POD5 | 1 (10) |
| POD6 | 3 (30) |
| POD7 | 2 (20) |
| POD13 | 1 (10) |
| Not reported | 1 (10) |

Abbreviations: CAS: Cumulated Ambulation Score, POD: Postoperative day

**Table 2.**
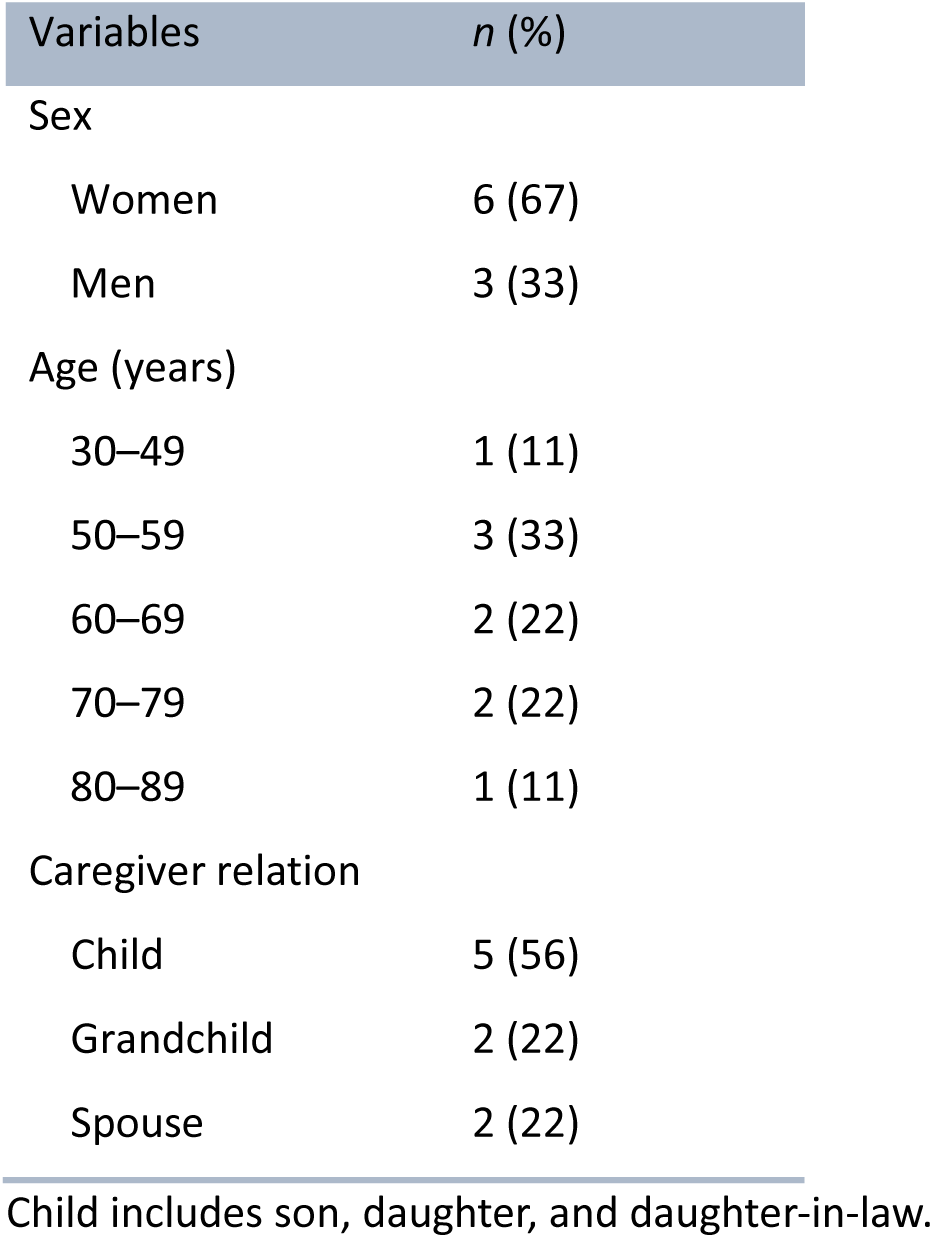
Description of caregivers (n=9).

| Variables | n (%) |
| --- | --- |
| Sex |  |
| Women | 6 (67) |
| Men | 3 (33) |
| Age (years) |  |
| 30–49 | 1 (11) |
| 50–59 | 3 (33) |
| 60–69 | 2 (22) |
| 70–79 | 2 (22) |
| 80–89 | 1 (11) |
| Caregiver relation |  |
| Child | 5 (56) |
| Grandchild | 2 (22) |
| Spouse | 2 (22) |

Child includes son, daughter, and daughter-in-law.

The thematic analysis revealed five overarching interrelated themes capturing how physical, psychological, communicative, structural, and organizational factors influenced patients’ and caregivers’ experiences of mobility during hospitalization after hip fracture. The themes were 1) Body and mind in transition; 2) Communication as a prerequisite for safety and mobility; 3) Structural barriers and ambiguities in responsibility; 4) The physical environment and ward culture; and 5) Mobility as preparation for life after discharge, shown in table 3.

**Table 3.** Overview of themes with subthemes.

| Themes | Subthemes |
| --- | --- |
| Body and mind in transition | - Physical decline and loss of autonomy<br>- Emotional vulnerability and motivation to regain autonomy and control |
| Communication as a prerequisite for safety and mobility | - Information, involvement, and shared understanding |
|  | - The tone of communication and relational treatment |
| Structural barriers and ambiguities in responsibility | - Resource constraints<br>- Unclear distribution of responsibilities |
| The physical environment and ward culture | - Environmental barriers<br>- Culture of inactivity |
| Mobility as preparation for life after discharge | - The need for individualized and meaningful mobility interventions<br>- Expectations and planning |

### Presentation of themes and subthemes

The themes reflect different analytical levels, ranging from more descriptive accounts of organizational and contextual conditions to more interpretive themes capturing participants’ lived experiences and how they interpret these experiences. This variation is consistent with the reflexive thematic analysis approach.

#### Body and mind in transition

This theme captures how hip fracture was experienced as a sudden transition involving intertwined physical and psychological changes that shaped engagement in mobility during hospitalization. Participants described an abrupt shift from independence to dependency, where bodily limitations, pain, and fatigue were closely linked to emotional vulnerability and concerns about future autonomy, positioning mobility as an uncertain and effortful practice rather than an activity taken for granted. Recovery was experienced as a dynamic process in which physical decline and emotional responses influenced motivation to move, with mobility continuously negotiated in relation to both bodily capacity and emotional engagement, and even small gains in mobility holding significant meaning.

##### Physical decline and loss of autonomy

For participants, a hip fracture represented a profound turning point in physical capability, suddenly characterizing previously simple activities difficult or impossible. The loss of strength, mobility, and stamina was described as deeply unsettling and disempowering, transforming mobility from a routine activity into a demanding and often uncertain effort. Many patients reported how even basic activities, such as getting out of bed, walking, or going to the bathroom or a chair, became significant challenges, thereby transforming everyday movement into effortful and uncertain tasks requiring support and attention. Pain and fatigue were pervasive barriers, constraining when and how patients felt able to engage in mobility.

> *“I was so tired that I could barely stand. I thought, now I’m going to fall.”* (Susan, in her early 70s)

Persistent pain compounded the sense of limitation, making movement both physically and emotionally demanding.

> *“It’s also because of the pain. I can’t go anywhere anymore, it just hurts.”* (Dorothy, in her early 80s)

Other symptoms, such as dizziness or side effects of medication, further restricted participation in rehabilitation.

> *“I get dizzy, and then I don’t dare to get up.”* (John, in his early 70s)

Physical dependence on others was described as a striking loss of autonomy and often accompanied by feelings of frustration or embarrassment, redefining mobility as a relational practice reliant on assistance rather than individual capacity.

> *“I couldn’t do it myself. So, I had to get some help to get around and to sit down and things like that.”* (John, in his early 70s)

Nevertheless, even small improvements, such as the ability to get out of bed or stand up, were described as important steps towards regaining independence and autonomy, illustrating how small gains accumulated and reshaped engagement in mobility over time.

> *“I was so happy yesterday when she brought that walker, so I could stand up by myself and go to the toilet and such. Instead of sitting and peeing in that bottle. I was given my independence back, you could say.”* (Robert, in his mid 80s)

Participants described how their altered bodies led to a reevaluation of self-reliance and the value of small, practical victories during recovery, redefining mobility as something that required constant adjustment and awareness.

##### Emotional vulnerability and motivation to regain autonomy and control

Emotional responses to hip fracture were closely intertwined with participants’ experiences of mobility during hospitalization, shaping how patients approached, avoided, or persisted with movement. The sudden dependency and loss of control led many to an immediate experience of vulnerability, insecurity, sadness, frustration, and sometimes hopelessness, which could discourage engagement in mobility or make movement feel risky and overwhelming. For many being unable to manage personal needs independently was not only frustrating but also emotionally distressing, undermining their identity as self-sufficient.

> *“It makes me sad. Yes, it does. Because I’ve always been someone who doesn’t want to be a burden. I’ve always managed on my own. I’ve always been self-reliant. And then you get put in your place when you have to ask someone for help. I don’t like that.”* (Joan, in her late 80s)

These experiences were sometimes accompanied by despair or a sense of resignation.

> *“You know, it’s terrible to say, but I wish I hadn’t woken up after my fall.”* (Joan, in her late 80s)

Yet, among these difficulties, the desire to return to a previous level of functioning and to regain autonomy persisted as a powerful motivator. Mobility was therefore not merely a physical task but a symbolic pathway toward regaining control, dignity, and identity, giving everyday movements meaning beyond their functional purpose. The fear of further decline, such as becoming permanently dependent, generated anxiety but also forced some patients to persist with rehabilitation, positioning mobility as both a source of distress and a necessary strategy to prevent further loss.

> *“I’m afraid that my body won’t cooperate if I stop moving.”* (Linda, in her late 60s)

For others, the limited time they felt remained in their lives was an important factor for recovery.

> *“The time I have left, I want to live.”* (William, in his late 80s)

Caregivers too recognized how closely the ability to move, and psychological well-being were intertwined, fearing that the loss of independence could undermine their loved ones’ will to live.

> *“She would lose her zest for life if she couldn’t get up and walk… That would be the end of her life. I think she would lose her will to live.”* (Sarah, in her early 50s)

Some participants found meaning and hope in even modest improvements.

> *“Now it works so much better, now I can do almost everything myself. Then I feel so much better, I think it’s going well.”* (John, in his early 70s)

These narratives revealed a complex negotiation between loss and adaptation, where mobility during hospitalization became both a source of emotional vulnerability and a means of restoring autonomy. Mobility emerged as a dynamic practice influenced by bodily limitations, emotional responses, and relational dependencies, with even small movements carrying profound psychological meaning and shaping how patients engaged in rehabilitation and everyday mobility practices.

#### Communication as a prerequisite for safety and mobility

Communication played a central role in shaping how patients and caregivers interpreted expectations regarding mobility, assessed whether movement was safe, and decided whether to initiate or postpone movement, thereby shaping mobility as a relationally mediated practice rather than an individual decision. The tone, clarity, and relational quality of healthcare professionals’ communication strongly influenced patients’ sense of safety, understanding, and involvement. Beyond conveying information, communication functions as a relational practice through which patients and caregivers feel seen, heard, and included in their care during a vulnerable period. Clear and attentive communication supported engagement in mobility, whereas inadequate or impersonal communication created uncertainty, passivity, and feelings of abandonment.

##### Information, involvement, and shared understanding

Participants consistently emphasized that efficient communication with healthcare professionals was essential to feeling secure, respected, and involved in mobility routines during hospitalization. When information about procedures, expectations, or progress was lacking or inconsistent, patients and caregivers described a state of confusion and discomfort. Not knowing what to expect or feeling excluded from important decisions, generated anxiety and a sense of disempowerment.

> *“It’s always… We do this. Oh no, we don’t anyway. Then we do it.”* (Richard, in his early 70s)

For some caregivers, the absence of structured and sufficient communication resulted in feeling unable to adequately support the patient and caregivers often assumed the role of advocates or informal coordinators, attempting to fill gaps in the information.

> *“I’m missing: ‘In order to return home or be discharged, one must be able to stand up independently or use the toilet alone.’ I don’t know what the goals are for an older woman in her 90s.”* (Carol, in her early 70s)

When expectations and mobility goals were unclear, patients and caregivers were uncertain about what was expected of them physically, which often led to passivity or hesitation in initiating movement. In such situations, mobility became something to wait for rather than to initiate, as patients hesitated to move without explicit encouragement or confirmation from staff, positioning patients as dependent on external confirmation before engaging in movement.

Caregivers depended on healthcare professionals’ information to support the patient, but when information was lacking or unclear, it led to frustration and limited caregivers’ ability to actively support patients’ engagement in mobility.

*“But I don’t want to cross the line and tell you how to do your job. But I thought it might have been a bit better, perhaps.”* (Chloe, in her early 30s)

Conversely, when healthcare professionals communicated clearly and worked collaboratively, caregivers and patients felt safer and more able to understand and participate in their care.

> *“You feel safer when they speak properly to you and explain what’s happening.”* (Patricia, in her early 70s)

Feeling safe and respected strengthened patients’ confidence to stand, walk, or practice daily activities, supporting more active engagement in mobility. In contrast, uncertainty or relational distance led some to remain seated or in bed despite the desire to move, thereby reinforcing passivity and delaying mobility. Harsh or impersonal communication undermined their sense of safety and reduced engagement in mobility-related activities. Communication failures, such as being left alone without access to a cord to call for help or not receiving essential information, further intensified feelings of helplessness and isolation, making patients hesitant to initiate mobility independently.

> *“I was left without a call cord. I couldn’t reach it and had to wait.”* (Margaret, in her early 80s)

##### The tone of communication and relational treatment

The tone and relational quality of interactions with healthcare professionals, as well as the overall atmosphere in the department, shaped patients’ sense of dignity, safety, and emotional security in relation to getting out of bed and moving. For some, interactions felt supportive and respectful; for others, they were experienced as abrupt or impersonal. These relational dynamics influenced not only emotional wellbeing but also how patients interpreted whether it was appropriate or safe to engage in mobility during hospitalization.

> *“The tone - if you only knew how harshly they spoke to her, both last night and this morning. It was a poor experience.”* (Patricia, in her early 70s)

While some described the tone as friendly, others felt spoken down to and ignored. Caregivers also note that the atmosphere varies depending on which staff member they interact with. Others shared experiences of witnessing or intervening in situations where fellow patients were treated without compassion or patience.

> *“I lay there crying. Even now I am touched by it. She was in her late 80s. I hobbled over to her and helped her. […] The worst experience has been hearing that tone used with such a dear old lady. […] She so much wants to be well and to have her normal life back. But instead, it’s just, ‘then you have to practice,’ bang.”* (Patricia, in her early 70s)

Overall, communication functioned as a relational and practical precondition for mobility. Clarity, relational attentiveness, and shared understanding enabled patients to interpret mobility as safe, expected, and legitimate, thereby supporting engagement in mobility. In contrast, uncertainty or relational distance created hesitation, dependency, and reduced initiative. Communication thus operated not merely as an exchange of information but as a determinant of how mobility-related activities were understood, initiated, and sustained, within everyday hospital care, shaping whether patients engaged actively or remained dependent and hesitant within mobility practices.

#### Structural barriers and ambiguities in responsibility

This theme captures how organizational structures and allocation of responsibility framed patients’ and caregivers’ experiences of mobility during hospitalization. Rather than focusing on individual motivation or physical capability, participants described how staffing levels, workflows, and unclear responsibility for mobility influenced whether movement was initiated, supported, or postponed, thereby structuring mobility as an organizationally dependent practice rather than an individually driven activity. Participants consistently pointed to system-level constraints that limited access to assistance, created uncertainty, and affected both safety and motivation to move.

##### Resource constraints

The organization and staffing of the department had a significant influence on perceived quality and safety of care during hospitalization, as well as on opportunities for mobility. Participants described insufficient resources, particularly staff shortages, as a major obstacle to mobility and rehabilitation. The experience of being forgotten led to frustration and decreased motivation for movement. Both patients and caregivers repeatedly cited the lack of available personnel, particularly physiotherapists during weekends and evenings, as a barrier to adequate mobility and care.

> *“I think maybe there should be more sessions. I know there aren’t many on weekends, and physiotherapists don’t work then. The only thing she [patient] did on the weekend was go to the toilet and sit for a while. Otherwise, she hasn’t walked because there weren’t enough staff.”* (Chloe, in her early 30s)

Several participants described having to request help multiple times, only to be left waiting or not receiving assistance at all. These experiences fostered feelings of helplessness and vulnerability, as the absence of predictable routines made the environment feel insecure and unreliable. Beyond formal rehabilitation activities, participants emphasized that assistance with basic personal needs, such as assistance with toileting, was closely intertwined with mobility. Accessing the toilet required standing, transferring, and walking, and therefore depended on staff availability and perceived responsiveness. Delays in receiving assistance with toileting were experienced as both physically uncomfortable and deeply undignified.

> *“I was once left sitting after a bowel movement and only wiped with one piece. I wasn’t aware because I was still numb in my leg, but I had to wait until the morning to remove the rest of the feces. I can’t understand how this happens.”* (Margaret, in her early 80s)

When help was delayed or unpredictable, patients described becoming hesitant to ask for assistance or to initiate movement. Mobility became associated with insecurity and dependency rather than progress, and movement was often postponed avoiding burdening staff. In this way, everyday mobility was experienced as contingent on staff availability rather than on patients’ readiness or rehabilitation needs, thereby limiting patients’ autonomy and shaping mobility as externally regulated.

#### Unclear distribution of responsibilities

In addition to resource constraints, patients and caregivers described considerable uncertainty regarding who was responsible for initiating and supporting mobility. While many patients expressed willingness to participate actively in their rehabilitation, they were often unsure whether mobility was their own responsibility or that of the staff. This ambiguity affected both confidence and willingness to move, positioning mobility as uncertain and potentially unsafe to initiate independently.

> *“I’m afraid of being forgotten, so I don’t get up.”* (Linda, in her late 60s)

This fear contributed to reduced initiation of movement and reinforced passivity. Caregivers similarly described difficulty identifying who held responsibility for different aspects of care, including mobility and medication. Inconsistent communication among staff further intensified this uncertainty.

> *“Sometimes, when she was supposed to get her blood thinner, the nurse didn’t know if she should have it and had to ask someone else. It made me wonder - shouldn’t you know? Should I know?”* (Chloe, in her early 30s)

This lack of clarity extended to the division of roles among different staff groups, leaving caregivers uncertain about whom they were interacting with.

> *“I don’t know if I’m speaking to a cleaner or a nurse.”* (George, in his early 80s)

When responsibility for mobility was unclear, patients described receiving insufficient encouragement or assistance to maintain independence, resulting in mobility being inconsistently supported and often deprioritized in daily practice.

> *“If there isn’t time, they don’t even ask if I want to get up from the chair.”* (Margaret, in her early 80s)

Over time, some patients stopped requesting help altogether.

> *“I ended up giving up asking, because I got tired of always having to request the same things.”* (Jennifer, in her early 50s)

Despite some positive encounters with motivated and professional staff, overall, these experiences demonstrated how unclear roles and responsibility for mobility created insecurity and hesitation, often resulting in unmet needs and reduced participation in mobility during hospitalization, where mobility was not embedded as a shared and continuous responsibility, but instead remained fragmented and inconsistently enacted in practice.

#### The physical environment and ward culture

Beyond organizational structures and staffing conditions, the physical environment and ward culture shaped implicit expectations about mobility, indicating whether movement was encouraged, expected, or deprioritized in everyday practice.

##### Environmental barriers

Participants described the physical surroundings of the department uninspiring and poorly suited to support movement or social interaction. Many were unaware of opportunities to move beyond their rooms, and access to walking areas or rehabilitation spaces was often limited or not clearly visible. This lack of visibility and accessibility reduced the likelihood that patients would initiate movement independently, reinforcing reliance on staff-led activities.

> *“I think many would like to use the space if it wasn’t just a storeroom. That can be a bit disappointing.”* (Linda, in her late 60s)

Several caregivers also noted that suitable assistive devices were unavailable, unused, or insufficiently prepared for patients.

> *“She hasn’t even tried her crutches. I don’t think they’ve even adjusted the size, or the height, as far as I know.”* (Richard, in his early 70s)

In addition, sensory aspects of the environments, such as noise and lighting, were described as draining energy and reducing motivation for activity.

> *“All the noise and activity make me so tired. I’m just used to things being quiet and peaceful.”* (Joan, in her late 80s)

Together, these environmental features were closely intertwined with a broader ward culture in which inactivity appeared normalized, positioning mobility as optional rather than expected and limiting patients’ opportunities to engage in movement beyond structured sessions.

##### Culture of inactivity

Alongside environmental barriers, patients and caregivers described a prevailing culture of the department in which passivity was normalized and movement was deprioritized, framing mobility as something contingent on scheduled interventions rather than an integral part of everyday care. Outside of scheduled physiotherapy sessions, there were few prompts or signals encouraging activity, and remaining in bed or seated was often experienced as the default expectation, reinforcing inactivity as the socially accepted norm within the ward.

> *“You’re just put in a chair and sit there waiting.”* (John, in his early 70s)

Some patients expressed frustration that their desire to remain active and continue everyday life was not reflected in ward practices.

> *“I want to get started, I want to continue my previous activities and life.”* (Barbara, in her early 80s)

Caregivers confirmed these observations, particularly during weekends or off-hours, when opportunities for movement appeared even more limited.

> *“She hasn’t really walked, except for going to the bathroom, because there aren’t many around, especially not on weekends.”* (Chloe, in her early 30s)

Together, these accounts illustrated how both the environments and the cultural aspects of the department functioned as barriers to meaningful mobility and rehabilitation, shaping mobility as a constrained and externally regulated practice rather than a self-initiated and integrated part of daily life.

#### Mobility as preparation for life after discharge

While the above themes describe barriers and constraints during hospitalization, this theme highlights how patients and caregivers orientated mobility toward life after *discharge*. Participants framed mobility not as an isolated in-hospital activity, but as a prerequisite for returning home, maintaining independence, and resuming everyday life.

##### The need for individualized and meaningful mobility interventions

Both patients and their caregivers strongly articulated the need for mobility interventions that were individualized, practical and meaningful to the patient’s daily life and rehabilitation goals. Participants highlighted the importance of early, proactive support to initiate mobility, as well as the essential role of staff in providing hands-on guidance and consistent follow-up. For many, simply receiving written instructions was perceived as inadequate without the structure and encouragement provided by healthcare professionals.

> *“Basically, I want more support and help to get out. It’s absolutely essential to walk, so you can get back to everyday life.”* (Margaret, in her early 80s)

Caregivers similarly recognized the value of active and continuous engagement by staff, noting that progress often depended on timely and attentive facilitation.

There was a pronounced desire for interventions that reflected the patient’s usual pre-fracture activities, routines and personal preferences, thereby making the rehabilitation process both relevant and motivating, increasing patients’ engagement in mobility and supporting more active participation in everyday mobility practices.

> *“It’s important that she can get up and sit down. She’s always been someone who got up and moved around, and now she’s very aware of how crucial that is.”* (Carol, in her early 70s)

Both patients and caregivers were generally critical of generic unsupervised exercise programs and underscored the need for individualized plans, regular monitoring, and clear feedback on progress.

> *“Instead of just handing over a piece of paper with exercises, there should be follow-up to make sure they’re being done.”* (Jennifer, in her early 50s)

Without such support, mobility risked becoming fragmented and inconsistently performed, rather than integrated into daily routines. Additionally, concerns were raised regarding the long-term implications of not regaining function, such as loss of independence and diminished ability to return home or participate in valued social activities. This future-oriented concern positioned mobility as essential work during hospitalization, rather than optional activity.

> *“If she couldn’t walk, I couldn’t see how she could manage at home… It would mean going from a completely independent mother to someone I wouldn’t know how to help.”* (Sarah, in her early 50s)

##### Expectations and planning

The recurring point from patients and caregivers was the need for clear, individualized, and proactive support and plan to guide mobility and rehabilitation and facilitate the transition home, which structured how and when patients engaged in mobility during hospitalization. There was a strong desire for structured assistance, more frequent engagement in mobility and a well-defined rehabilitation plan tailored to patients’ needs.

> *“It’s much better to have a schedule, a plan, maybe for a whole group or a ward, since everyone is here because of a hip injury.”* (Dorothy, in her early 80s)

Several participants emphasize the importance of timely, consistent, and transparent information and support for physiotherapists or staff, without which patients might remain passive and less mobile.

> *“If the physiotherapist hadn’t come and got her out of bed on the first day, she would still just be lying there.”* (David, in his early 60s)

This highlights how mobility was often dependent on external initiation, rather than self-driven activity. Patients also expressed the need for assistance and guidance in everyday activities as prolonged inactivity was perceived as physically demanding, indicating that inactivity itself shaped patients’ bodily capacity and readiness to move.

> *“I would get more help… just sitting or lying in the same position is hard.”* (Margaret, in her early 80s)

Both patients and caregivers valued the role of mobility in regaining previous function and maintaining a meaningful daily life. This was seen in references to personal goals, such as returning to hobbies and independent activities.

> *“It’s super important, because she’s such an active person and wants to get back to bowling, gardening, and all the things she used to do.”* (Jennifer, in her early 50s)

Participants appreciated movements that were frequent and embedded within the rhythms of everyday life, opportunities to walk or engage in activities, as well as more social or collaborative rehabilitation environments.

> *“It would be nice if there was more time to walk together with someone.”* (John, in his early 70s)

These preferences suggest that mobility was more likely to be sustained when embedded in social interaction and everyday routines. Finally, there was a need for transparency about what to expect and how progress in recovery would be measured, both in-hospital and post-discharge.

> *“Where is the follow-up, the clear agreement on what is expected in her situation?”* (Sarah, in her early 50s)

Collectively, these perspectives revealed a gap between the participants’ expectations and the current mobility interventions and planning, where mobility was not consistently supported as a structured, meaningful, and future-oriented practice, but instead occurred sporadically with limited integration into everyday care.

Taken together, the findings demonstrate that mobility during hospitalization was shaped by a complex interaction of physical limitations, organizational structures, communication practices, cultural norms, and future-oriented expectations, influencing whether movement was encouraged, postponed, or abandoned.

## DISCUSSION

This study explored how patients and caregivers experience mobility during hospitalization after hip fracture surgery. Our findings suggest that mobility during hospitalization was experienced not merely as a physical task but as a socially shaped and negotiated practice embedded in everyday hospital routines. These interactions appeared to shape patients’ engagement in mobility by providing or limiting cues about whether movement was safe, expected, and appropriate, thereby influencing whether mobility was initiated or postponed. This understanding aligns with Kirk et al. (67), who describe mobility during hospitalization as a socially and organizationally negotiated practice shaped by everyday interactions, routines, and implicit expectations rather than individual capacity alone.

While the broader rehabilitation literature often distinguishes between mobilization, physical activity, and exercise therapy, participants in this study did not differentiate between these concepts. Instead, they described mobility as a unified, functional experience centered on getting out of bed and moving. This suggests a potential gap between clinical terminology and patient experience and supports the use of mobility as the central analytic concept in this study. From this perspective, our findings extend existing understandings of post-operative mobility by conceptualizing mobility not only as an individual movement but as a socially shaped and negotiated practice embedded in hospital routines. By foregrounding relational, communicative, and contextual dimensions, the study contributes to a more nuanced understanding of how mobility is enacted in clinical settings.

A central contribution of this study is the conceptualization of mobility as a socially negotiated practice rather than a purely physical task. Patients’ engagement in mobility appeared to depend not only on physical capacity, but also on how movement was legitimized through communication, staff availability, and organizational routines. Unclear responsibility and inconsistent support constrained patients’ willingness to initiate movement, positioning mobility as contingent on external validation rather than individual readiness. This highlights how mobility is shaped by relational and organizational conditions, extending existing literature beyond primarily individual and logistical explanations.

Our findings extend previous research by showing that barriers to mobility are not only practical or logistical but also relational and contextual (6,32,33). Participants described how feelings of safety, dignity, and clarity about expectations influenced their willingness to move during hospitalization. From a rehabilitation perspective, this underscores that mobility after hip fracture cannot be understood or addressed in isolation from the social and organizational environment in which it takes place. Similar relational barriers have been identified in staff-centered studies describing time pressure, fragmented workflows, and limited interprofessional coordination (68,69). Our study adds the patient and caregiver perspective by illustrating how these system-level conditions are experienced as uncertainty, passivity, and hesitation to initiate movement.

Consistent with earlier qualitative studies (5,6), patients described hip fracture as a sudden and profound disruption of both bodily function and independence, which shaped their engagement in mobility during hospitalization. This reflects broader qualitative evidence indicating that hip fracture is experienced as a life-altering event marked by psychosocial distress, including loss of identity, fear of dependency, and uncertainty about the future (8). Pain, fatigue, and fear of falling limited physical capacity, while emotional vulnerability and dependency further shaped engagement in mobility. For several participants, relying on others for basic activities challenged deeply held values of independence and self-reliance, sometimes leading to withdrawal and passivity. However, our findings also revealed a persistent motivation to regain autonomy. Even small functional gains were experienced as meaningful and motivating, and these small gains appeared to accumulate over time, indicating that mobility served not only a physical but also a symbolic function in recovery. In this sense, mobility became a way of restoring control, reflecting previous findings that patients perceive recovery as a personal journey requiring psychological readiness and contextual support (9). Our findings specify how such readiness is shaped in the acute hospital setting, where encouragement, clear information, and opportunities to make decisions about movement appeared to shape patients’ motivation by increasing their confidence and sense of control, thereby supporting more active engagement in mobility. This aligns with research emphasizing that rehabilitation after hip fracture involves renegotiating identity and agency (4,29). These findings underline the importance of addressing emotional as well as physical dimensions of mobility during acute hospitalization, rather than focusing narrowly on functional milestones.

Communication emerged as a central mechanism through which mobility was negotiated during hospitalization. Participants described how clear, respectful, and relational communication fostered feelings of safety and involvement, whereas inconsistent or impersonal communication generated uncertainty and disengagement. Communication appeared to shape whether movement was safe, appropriate, and expected, thereby influencing patients’ willingness to initiate movement. These findings resonate with previous studies showing that communication influences patients’ confidence and participation in rehabilitation (10,35). The present study adds nuance by demonstrating how communication affected not only patients’ engagement in mobility, but also caregivers’ ability to support and encourage mobility. Importantly, caregivers were not passive observers but appeared to function as active mediators of mobility by interpreting information, encouraging movement, and compensating for limited communication and coordination from staff. When goals, expectations, and progress were unclear, caregivers described difficulty fulfilling this role, sometimes assuming informal coordinating functions. This highlights the relational nature of rehabilitation and supports emerging literature positioning caregivers as a bridge between hospital and home, often carrying substantial responsibility for post-discharge rehabilitation despite limited involvement during hospitalization (35,36). Indeed, recent Scandinavian evidence shows that up to 90% of older patients receive help from caregivers after hip fracture, with a median of 32 hours support during the first weeks following discharge (70), underscoring the central role caregivers play in supporting recovery after discharge. Difficulties in communication also aligned with staff-centered research identifying fragmented interdisciplinary coordination and lack of shared understanding as organizational barriers to effective hip fracture care (32). While such study describes these challenges from a professional perspective, the present findings illustrate how fragmented communication is experienced by patients and caregivers as inconsistency, lack of clarity, which reduced patients’ and caregivers’ involvement by limiting their understanding of expectations and thereby constraining engagement in mobility. Moreover, our findings resonate with research on communication with older adults, showing that overly simplified, directive, or patronizing communication may undermine older adults’ sense of competence and agency (71). Although specific communicative styles were not systematically assessed, participants’ descriptions of harsh tone, lack of explanation, and feeling talked down to suggest that ‘how’ communication is delivered, not only ‘what’ is said, may influence motivation and willingness to engage in mobility.

Participants also described how organizational structures constrained opportunities for mobility during hospitalization. Limited staffing and unclear responsibility for initiating and supporting mobility created hesitation about when movement was expected, encouraged, or safe. This uncertainty may partly reflect a lack of shared language and common understanding of mobility-related tasks, as previously identified in staff-centered ethnographic research, where mobilization, physical activity, and exercise were interpreted and enacted differently across professional groups (58). While such distinctions are known in rehabilitation literature, participants in this study did not differentiate between these concepts, instead describing mobility as a unified, functional experience centered on getting out of bed and moving. This mismatch between professional terminology and patient experience may further contribute to uncertainty about roles and expectations by obscuring what mobility entails in practice, thereby reducing patients’ and caregivers’ ability to engage actively in movement. These findings are consistent with studies identifying organizational barriers to early mobility after hip fracture (28,32), but they extend this literature by illustrating how such barriers are experienced at the patient and caregiver level. Rather than being perceived as abstract system issues, organizational constraints were experienced as insecurity, and fear of being forgotten, which appeared to reduce patients’ willingness to request assistance and thereby limit opportunities to engage in mobility. This highlights how mobility is not solely an individual capability, but a practice dependent on organizational conditions and distributed responsibility.

In addition to organizational structures, the physical environment and ward culture played a significant role in shaping expectations about movement. Uninspiring surroundings and limited environmental cues appeared to normalize inactivity outside formal therapy. This normalization of passivity suggests that inactivity was not merely a consequence of individual fatigue or lack of motivation, but part of an institutionalized culture in which remaining in bed or seated was perceived as the default and safest option, not only in terms of physical risk, but also because patients felt they needed to remain available for ward rounds, information, or assistance from staff. Similar patterns have been described in staff-centered ethnographic research, where inactivity became the “natural” state during hospitalization, while mobilization required explicit justification and legitimation (58). From this perspective, mobility is not simply an individual behavior but a practice that needs to be socially sanctioned within the ward. When cues, routines, and professional interactions implicitly signal that rest is the norm, patients may hesitate to move even when physically able or motivated. This finding aligns with literature on hospital-induced immobility, which highlights how environmental and cultural factors contribute to sedentary behavior during hospitalization (27,59). The present study adds a patient-centered perspective by illustrating how such environments communicate implicit messages about what is appropriate or expected behavior. When movement is not visibly supported or encouraged, patients may interpret inactivity as the safest or most acceptable option in response to limited environmental cues and support for movement, thereby reinforcing passivity even when physically able to move. Rehabilitation efforts aimed at increasing mobility may therefore benefit from addressing not only individual behavior but also the environmental and cultural context of care.

A key finding to our study was that patients and caregivers consistently framed mobility during hospitalization in relation to returning home and resuming everyday life. Mobility was therefore valued less as an isolated activity and more as preparation for independence after discharge. This emphasis on independence aligns with previous qualitative evidence showing that the prospect of long-term dependency may be experienced as profoundly distressing, with some patients expressing that a life marked by severe dependency may no longer feel worth living (8). Participants emphasized the need for individualized support, clear goals, and structured planning to make mobility meaningful and motivating. This forward-looking orientation has been described in previous studies (5,72), but our findings highlight how gaps in in-hospital mobility support and communication can undermine confidence in managing at home. From a rehabilitation perspective, aligning early mobility with patients’ everyday lives and post-discharge trajectories may enhance engagement during hospitalization by increasing the perceived relevance of movement for future independence.

Although initiatives such as Network for the Physically Active Hospital (NEFAH) (73) and mobility-promoting interventions have highlighted the importance of structured pathways for mobility (37,74), our findings suggest that mobility during hospitalization is also shaped by relational and contextual factors within everyday ward practice. From an intervention perspective, efforts to increase early mobility must therefore attend not only to what patients are expected to do, but also how mobility is communicated legitimized, and socially supported within ward routines. Without attention to these relational and contextual components, even well-designed interventions may fail to translate into meaningful movement for patients. This gap between evidence, policy and everyday practice has been described in implementation research as a knowing-doing gap, where established knowledge fails to be consistently enacted in clinical settings due to organizational constraints, fragmented workflows, and competing priorities in clinical work (75,76). Similar challenges have been identified in staff-centered research, showing how mobility is shaped by routines and responsibility structures rather than clinical intentions alone (32). From a patient and caregiver perspective, this knowing-doing gap materialized as prolonged inactivity, uncertainty about expectations, and reliance on chance encounters with staff to initiate mobility. While staff-centered studies primarily describe these challenges from a professional viewpoint, the present findings illustrate how they are experienced at the patient level as inconsistent encouragement, unclear expectations, and limited support for movement.

Taken together, this study contributes a patient- and caregiver-centered understanding of early mobility after hip fracture as a negotiated, relational, and context-dependent practice. It highlights how organizational, communicative, and cultural conditions shape not only opportunities for movement, but also patients’ confidence, motivation, and sense of legitimacy in mobility. These insights may inform the development and refinement of future mobility-supporting interventions by prioritizing patient and caregiver perspectives in intervention design and implementation.

### Strengths and limitations

A key strength of this study is the inclusion of both patients and caregivers, which provides a more comprehensive understanding of factors influencing mobility during hospitalization. The use of reflexive thematic analysis was valuable in this study, as it enabled identification of patterns of shared meaning across participants while allowing for interpretive engagement with how mobility was experienced and negotiated in everyday clinical practice. Conducting interviews during the acute hospital phase enabled capture of experiences while mobility was actively negotiated, offering insight into real-time barriers and facilitators as they unfolded in practice. Several limitations should be acknowledged. Interviewing participants during hospitalization may have constrained their ability to reflect on longer-term consequences and recovery trajectories beyond discharge. Patients with cognitive impairment were not included, which may limit transferability to the broader hip fracture population. However, caregivers of patients with cognitive impairment were included and were able to reflect on the mobility-related challenges experienced by these patients during hospitalization. Although this provides indirect insight into this vulnerable group, the absence of direct perspectives from cognitively impaired patients remains a limitation. The study was conducted at a single hospital context, and the findings are therefore context-specific, which may limit transferability to other settings. Future research could build on these findings by exploring patient and caregiver experiences longitudinally across the transition from hospital to home, to inform the development and implementation of mobility-supporting interventions.

## CONCLUSION

This study shows that mobility during hospitalization after hip fracture surgery extends beyond physical capacity and is more appropriately understood as a socially negotiated and context-dependent practice. Patients’ engagement in mobility was shaped by how movement was communicated, supported, and organized within everyday clinical routines, influencing whether mobility was initiated or postponed. From this perspective, mobility is not solely an individual task but depends on relational and organizational conditions that shape patients’ sense of safety, autonomy, and readiness to act. For patients and caregivers, mobility was closely linked to dignity and the possibility of returning to everyday life, highlighting its significance beyond functional recovery. These findings suggest that efforts to improve mobility during hospitalization should attend not only physical rehabilitation, but also to how mobility is legitimized and supported within clinical practice.

## Data Availability

Data are not available.

## ACKNOWLEDGMENTS

The authors thank all the participants in this study for their time and willingness to share their perspectives.

## IMPLICATIONS FOR REHABILITATION

- Patients’ engagement in mobility appears closely linked to perceptions of safety, dignity, and clarity of expectations.
- Mobility may be influenced by organizational conditions, including staffing, workflows, and distribution of responsibility.
- Communication and relational interactions seem central in shaping whether mobility is initiated, supported, or postponed.
- Linking in-hospital mobility to patients’ everyday lives and post-discharge goals appears meaningful for engagement in rehabilitation.

